# Development and validation of chest CT-based imaging biomarkers for early stage COVID-19 screening

**DOI:** 10.1101/2020.05.15.20103473

**Authors:** Xuanyu Mao, Xiao-Ping Liu, Miao Xiong, Xu Yang, Xiaoqing Jin, Zhiqiang Li, Shuang Zhou, Hang Chang

## Abstract

Coronavirus Disease 2019 (COVID-19) is currently a global pandemic, and the early screening of COVID-19 is one of the key factors for COVID-19 control and treatment. Here, we developed and validated chest CT-based imaging biomarkers for COVID-19 patient screening. We identified the vasculature-like signals from CT images and found that, compared to healthy and community acquired pneumonia (CAP) patients, the COVID-19 patients revealed significantly higher abundance of these signals. Furthermore, unsupervised feature learning leads to the discovery of clinical-relevant imaging biomarkers from the vasculature-like signals for accurate and sensitive COVID-19 screening that has been double-blindly validated in an independent hospital (sensitivity: 0.941, specificity: 0.904, AUC: 0.952). Our findings could open a new avenue to assist screening of COVID-19 patients.

## Introduction

Coronavirus Disease 2019 (COVID-19) is currently a global pandemic[1, 2]. More than one million cases of COVID-19 have currently been confirmed, but the peak in the number thrives through uncertainty [3]. China’s prevention and control experience in February showed that early detection, early diagnosis, early isolation, and early treatment are essential for the prevention and control of the entire COVID-19 epidemic. Nucleic acid detection based on the new coronavirus provides the most important tools for the diagnosis of pathogens and COVID-19. However, many challenges include: (1) COVID-19 belongs to a class of highly infectious diseases, and a considerable proportion of the patients have no obvious clinical symptoms during the onset of this disease [4]; (2) in the early stages of the disease, the amount of virus in the upper respiratory tract, including the nasopharynx, is very small under detection, which result in that some patients with suspected COVID-19 cannot be diagnosed, isolated, and treated because of negative nucleic acid test results; (3) due to the quality of the coronavirus nucleic acid detection kit is not uniform, the operation is not standardized and other factors, the nucleic acid test has obvious false negatives; and (4) due to the rapid outbreak of the epidemic in many regions around the world, there are obvious shortages of resources including nucleic acid detection kits in some countries, which also limits the clinical application of the nucleic acid detection.

Except for the coronavirus etiology, epidemiological contact history, and clinical symptoms, pulmonary imaging (especially chest computed tomography (CT) imaging) has very unique value for the diagnosis of COVID-19[5]. COVID-19 patients with early lung CT showed multiple interstitial changes in small patchy albums, which were evident in the extrapulmonary bands. As the disease progresses, the imaging of the disease can develop into multiple “ground-glass opacities” (GGO) and infiltrates in the lungs, and severe consolidation may occur in patients with severe disease [6]. However, the CT image of COVID-19 patients especially at early stage are similar to these from other common pneumonia patients, including H7N9 influenza virus pneumonia, mycoplasma pneumonia, chlamydial pneumonia and bacterial pneumonia [7]. In this study, we develop and validate chest CT-based imaging biomarkers for COVID-19 patient screening using artificial intelligence (or computer vision) methods, which will be of great significance to reduce the workload of clinicians and to assist in differential diagnosis of COVID-19 from other diseases. Our findings could open a new avenue to assist screening of COVID-19 patients.

## Material and Methods

### Data collection

The chest CT images in this case-control study were collected from Hubei Provincial Hospital of Traditional Chinese Medicine and Wuhan Third Hospital. The inclusion criteria for COVID-19 patients were: (1) patients were diagnosed and confirmed through nucleic acid test from January 2020 to March 2020; (2) patient were with mild or moderate disease status, where the severity was classified according to the Coronavirus Disease 2019 (COVID-19) diagnosis and treatment guideline (trial version 7) issued by the National Health Commission of the People’s Republic of China. Specifically, according to the guidelines, patients with mild disease status have mild clinical symptoms, and have no obvious pneumonia manifestations on chest CT images; and patients with moderate disease status have obvious respiratory symptoms such as fever and cough, and have obvious pneumonia manifestations on chest CT images. In addition, both patients with community acquired pneumonia (CAP) and healthy participants (with no obvious abnormalities in chest CT images) were randomly collected from aforementioned two hospitals and used as control group. The inclusion criteria for control group were: (1) patients who were diagnosed with lung infection on imaging and clinical basis few months before the onset of the epidemic; (2) patients without severe diseases of respiratory system, cardiovascular or cerebrovascular systems, (3) patients without mental illness or cognitive impairment. This study has been approved by the institutional review board (IRB) of both participating hospitals.

### Imaging Protocol for CT chest

Chest CT exams from Hubei Provincial Hospital of Traditional Chinese Medicine were performed with two different scanners: (1) GE Optima 660 CT (GE Healthcare, Milwaukee) and (2) uCT 530 (United imaging, Shanghai), with reconstruction thickness at 0.625 mm and 1 mm, respectively. While, CT exams from Wuhan Third Hospital were performed with GE Discovery CT750 HD (GE Healthcare, Milwaukee) with reconstruction thickness at 0.625mm.

### Vasculature-like Structure Enhancement

Vasculature-like signal is recognized and enhanced using an iterative tangential voting (ITV) approach [8] within pre-segmented lung regions in 3D, where ITV enforces the continuity and strength of local linear structures (i.e., vasculature-like structure) and the 3D lung segmentation is achieved via level-set method [9]. Specifically, ITV operates on CT image gradient information with sigma set to be 0.5 and 1.0 on training and validation cohorts, respectively, to accommodate the technical difference across hospitals.

### Imaging Biomarker Detection and Visualization for COVID-19 Screening

We developed an unsupervised feature learning pipeline based on Stacked Predictive Sparse Decomposition (Stacked PSD) [10] for the unsupervised discover of underlying 3D characteristics from the ‘vasculature-like signal’ space derived from CT-based raw images. Specifically, in this study, we used single network layer with 256 dictionary elements (i.e., signal patterns) at a fixed pattern size of 20×20×20 pixels and a fixed sampling rate of 100 3D patches per sample which were experimentally optimized. In the training cohort, 8 of 256 dictionary elements were identified to have significant correlation with COVID-19 with cutoff FDR value < 0.05 through cross-validation approach (training sample rate: 0.8; bootstrap 100 times). At last, these 8 significant dictionary elements as a set of imaging biomarkers were selected and utilized to build the random decision forests model for COVID-19 screening. A double-blind study was designed and implemented to validate this model in an independent hospital. Visualization of these imaging biomarkers was created in three-dimensional space using ITK-Snap (version 3.8.0), Python (version 3.7.0), Matplotlib (version 3.1.2), Blender (version 2.82) and Three.js (version r115 on GitHub). Snapshots of the three-dimensional visualization were used to generate two-dimensional visualization that overlays with the original CT slices.

### Comparison of the Performance between the 3D Imaging Biomarkers and Experienced Chest Radiologists

In order to validate the diagnostic performance of our pre-identified 3D imaging biomarkers, we invited two experienced chest radiologists to independently and blindly (blind to clinical data) assess the CT images in our validation cohort. These two chest radiologists have 8 and 10 years of clinical imaging diagnosis experience, respectively. And both of them have more than 2 months of intense and continuous diagnosis experience of COVID-19 in Wuhan, China. Sensitivity and specificity were utilized for performance comparison between the chest radiologists and our 3D imaging biomarkers.

### Statistical Analysis

The difference in the vasculature-like signals among different groups (COVID-19, CAP and healthy) was assessed by non-parametric test, and the association between signatures and COVID-19 by logistic regression. Principle component analysis (PCA) and heatmap cluster analysis were performed in R (version 3.6.1) and MATLAB (version 2012b), respectively. The screening performance was characterized with sensitivity, specificity and area under curve (AUC).

## Results

### Study Population Characteristics

In order to identify chest CT-based imaging biomarkers for COVID-19 patient screening, we conducted a case-control study in two hospitals together with artificial intelligence technologies in machine learning (Fig. 1). The population characteristics for the training and double-blind validation cohorts are summarized in Extended Data Table 1. A total of 321 participants were included in this case-control study. The cohort (n=116) from one hospital (Hospital A) served as training set, the cohort (n=205) from the other (Hospital B) as a double-blind validation set (Fig. 1). The median ages of participants in the training and validation cohorts were 42 (range: 14-76) and 58 (range; 19-89), respectively. There were 53 (45.7%) females and 63 (54.3%) males in the training cohort, while corresponding data were 110 (53.7%) and 95 (46.3%) in the validation cohort. The training cohort contained 47 (40.5%) COVID-19 patients, 20 (17.2%) healthy and 49 (42.2%) CAP patients, while the validation cohort had 153 (74.6%) COVID-19 patients, 15 (7.3%) healthy and 37 CAP (18%) patients.

**Figure 1.**
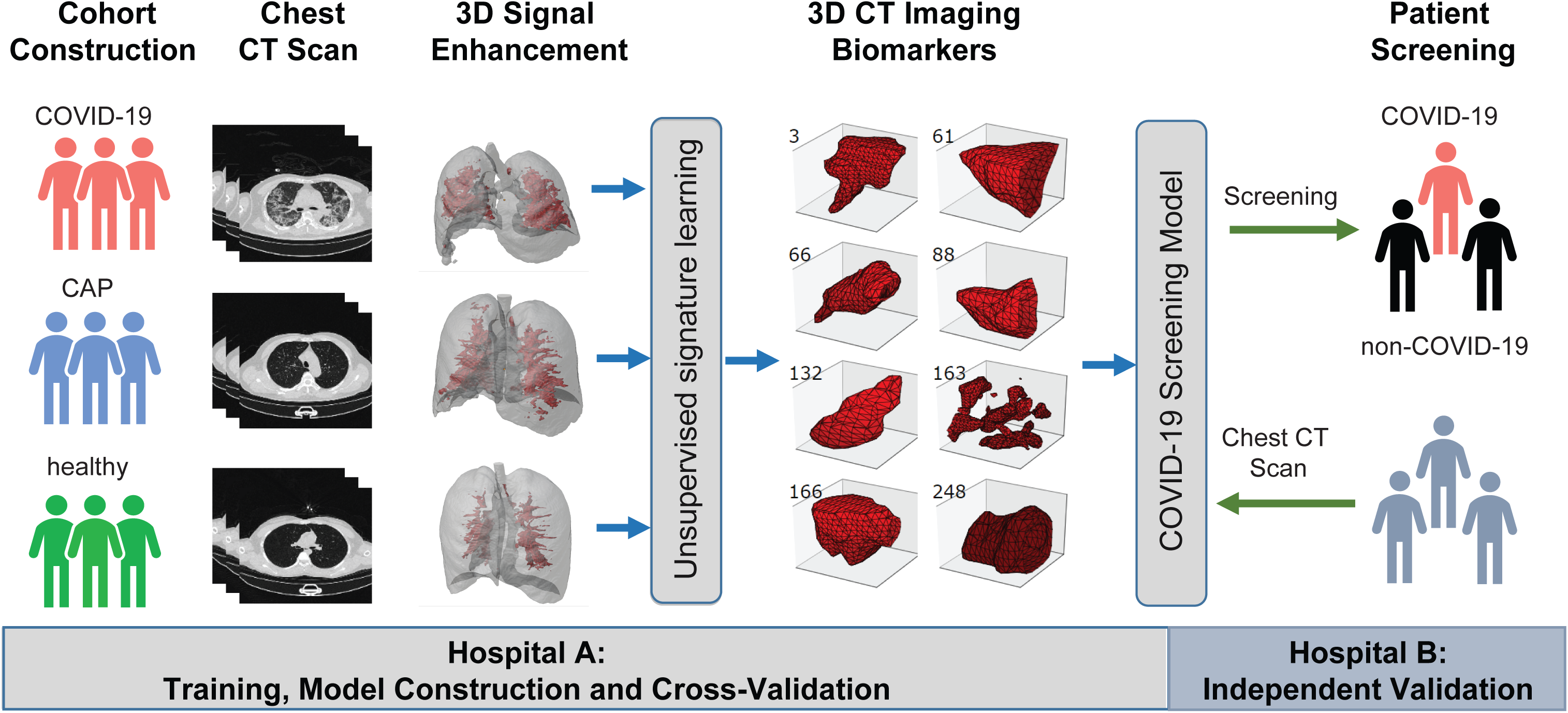
A graphic illustration of the study design. A case-control study was designed to identify chest CT-based imaging biomarkers for COVID-19 patient screening. Biomarker discovery and biomarker-based predictive model construction were conducted using the data from Hospital A (training cohort), which were validated in Hospital B (validation cohort) with the double-blind design.

### Vasculature-like Structure Enhancement

In our study, the vasculature-like structure was recognized and enhanced with iterative tangential voting in both training and validation cohorts as a pre-processing step. Interestingly in the training cohort, the mean vasculature-like signal reveals significant difference (p-value < 0.05) among healthy, CAP and COVID-19 patients (Fig. 2b). These findings are consistent with the observation of vascular changes in lung tissue from COVID-19 patients, including vascular congestion/enlargement, small vessels hyperplasia and vessel wall thickening[11, 12]. Furthermore, such distinction itself leads to remarkable differentiation between COVID-19 and non-COVID-19 groups in our training cohort (AUC=0.721, Extended Data Fig. 2, blue curve) with logistic regression approach. Altogether, those results encourage us to identify imaging biomarkers from the “vasculature-like signal” space to assist accurate screening of COVID-19.

**Figure 2.**
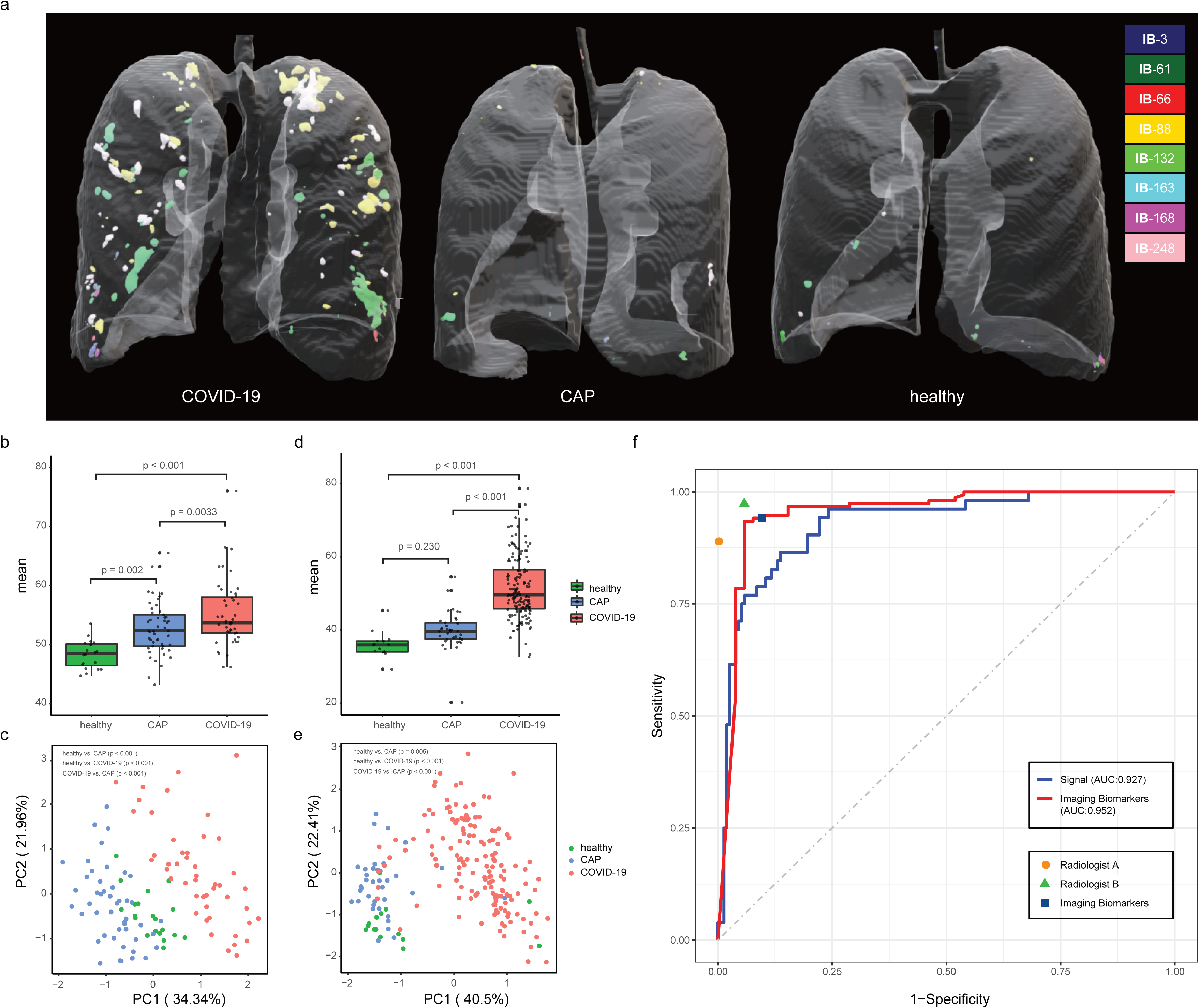
Chest CT-based imaging biomarkers highly predicts COVID-19. **a**. Representative examples for 3D multispectral imaging biomarker visualization in COVID-19, CAP and healthy samples **b**. The boxplot shows differences in the vasculature-like signals among healthy, community acquired pneumonia (CAP), and COVID-19 patients in the training cohort. The p-values were obtained by the non-parametric Mann-Whitney test. **c**. PCA of 8 imaging biomarkers in the training cohort. 20 healthy participants (green dots), 49 CAP patients (blue dots), and 47 COVID-19 patients (red dots). The p-values were obtained from permutational multivariate analysis of variance (PERMANOVA). **d**. The boxplot shows differences in the vasculature-like signals among healthy, community acquired pneumonia (CAP), and COVID-19 patients in the validation cohort. The p-values were obtained by the non-parametric Mann-Whitney test. **e**. PCA of 8 imaging biomarkers in the validation cohort. 15 healthy participants (green dots), 37 CAP patients (blue dots), and 153 COVID-19 patients (red dots). The p-values were obtained from permutational multivariate analysis of variance (PERMANOVA). **f**. Screening performance of signal-based model, imaging biomarker-based model, and two COVID-19 experienced radiologist on validation cohort.

### Imaging Biomarker Detection and COVID-19 Screening

Next, we applied Stacked PSD on the entire training cohort within the ‘vasculature-like signal’ space to acquire underlying characteristics for dictionary construction (details see method). 256 dictionary elements were learned and optimized from the entire training cohort. We found that 8 of 256 dictionary elements have significantly positive correlation with COVID-19 (FDR < 0.05, Extended Data Table 2 and Supplementary Table 1). These 8 COVID-19-relevant signatures (i.e., imaging biomarkers) are graphically presented (Fig. 1 3D CT Imaging Biomarkers panel). These biomarkers also allow the construction of full 3D multispectral staining in the entire lung region (Fig. 2a), which is further demonstrated in 3D animations (Supplementary Videos 1-3). The corresponding 2D multispectral staining into the CT image slices are also constructed (Supplementary Videos 4-6). The 8 imaging biomarkers clearly separate COVID-19 patients from others in the training cohort by PCA (Fig. 2c) and clustering (Extend Data Fig. 3a) analysis. Finally, we built a random decision forest model for COVID-19 screening based on these imaging biomarkers within the training cohort (AUC = 1.000, Extended Data Fig. 2, red curve), which will be tested in the validation cohort.

### Test of Imaging Biomarkers in Validation Cohort

Identical enhancement process was applied onto the validation cohort. Similar to training cohort, we observed the distinction of mean vasculature-like signal among different groups (i.e., COVID-19, CAP and healthy) (Fig. 2d). The logistic regression model pre-built in the training cohort on signal led to accurate prediction between COVID-19 patients and others in the validation cohort (AUC=0.927, Fig. 2f, blue curve). 8 pre-identified imaging biomarkers also clearly separate the COVID-19 patients from others in validation cohort (Fig. 2e, Extended Data Fig. 3b). Excitingly, we founded the pre-built random decision forest model based on pre-obtained biomarkers predict COVID-19 with high sensitivity (0.941), specificity (0.904), and AUC (0.952), which is competitive with two COVID-19 experienced chest radiologists (Fig. 2f).

## Discussion

In this study, we developed and validated 3D imaging biomarkers for COVID-19 screening based on chest CT images. Our quantitative evaluation suggests that, compared to healthy and CAP patients, COVID-19 patients may have significantly more vascular changes in lung tissue, including vascular congestion/enlargement, small vessels hyperplasia and vessel wall thickening[11, 12], which leads to the discovery of robust imaging biomarkers for COVID-19 screening. Our double-blind validation confirms the robustness and effectiveness of pre-identified imaging biomarkers in an independent hospital with high specificity (0.904) and sensitivity (0.941), which is competitive with two COVID-19 experienced chest radiologists.

The current COVID-19 epidemic is a world-wide threat. Specifically, in Europe and the United States, there are tens of thousands of new confirmed cases and suspected cases every day[13-16]. These sharply increased cases of COVID-19 are running out of medical resources in some countries to varying degrees and are paralyzing the medical system in some countries[17]. Therefore, rapid screening, diagnosis, isolation and treatment of COVID-19 patients are particularly important for the prevention and control of the epidemic. However, it is unfortunate that nucleic acid detection kits for COVID-19 have been in short supply in many countries. At the same time, due to improper operation, technical variations among different nucleic acid detection kits and many other reasons, the results of nucleic acid test have certain false negatives. On the other hand, CT examination has been proved to have unique advantages in the early screening and diagnosis of COVID-19[11, 18]. To discover new possibility for COVID-19 screening, this study utilized an unsupervised deep learning method to identify robust imaging biomarkers from chest CT scans for accurate COVID-19 screening. The major advantages of our imaging biomarkers reside in two folds as follows: (1) they provide robust, accurate and cost-effective COVID-19 screening, which can significantly alleviate the shortage of clinical resources, including both nucleic acid detection kids and experienced chest radiologists; and (2) they provide a non-invasive diagnostic tool that enables world-wide scalable practical applications. Furthermore, our imaging biomarkers may provide a new avenue for predicting COVID-19 patients’ prognosis and clinical outcome, which will be further investigated in our future research.

## Data Availability

The 3D imaging biomarkers and the pre-trained Stacked PSD model are available at http://bmihub.org/project/covid19-imaging-biomarkers. The raw chest CT cohorts involved in this study were not publicly available due to the DICOM metadata containing information that could compromise patient privacy/consent.

## Conflicts of Interest

The authors declare no conflicts of interest

## Author Contributions

J.M., H.C., M.X. and S.Z. designed the study; X.L., X.M., X.Y. and H.C. performed the analysis; J.M., H.C. and X.L. wrote the manuscripts; All authors revised the manuscript.

**Abbreviations**
COVID-19: Coronavirus Disease 2019
GGO: ground-glass opacities
CT: computed tomography
PSD: Predictive Sparse Decomposition
CAP: community acquired pneumonia
ITV: iterative tangential voting

## Data Availability

The Chest CT images involved in this study are available upon request and consideration by corresponding author(s) of this manuscript.

## Code Availability

Iterative Tangential Voting for vasculature-like structure enhancement is publicly available at http://bmihub.org/project/itv; and_the Stacked PSD for imaging biomarker detection is publicly available at http://bmihub.org/project/stackedpsd.

## Notes

### Competing Interest Statement

The authors have declared no competing interest.

### Funding Statement

no external funding was received

### Author Declarations

This study has been approved by the institutional review board (IRB) of both participating hospitals

## References

1. Velavan TP, Meyer CG: The COVID-19 epidemic. Trop Med Int Health 2020, 25(3):278–280.

2. Mian A, Khan S: Coronavirus: the spread of misinformation. BMC Med 2020, 18(1):89.

3. Kinross P, Suetens C, Gomes Dias J, Alexakis L, Wijermans A, Colzani E, Monnet DL, European Centre For Disease P, Control Ecdc Public Health Emergency T: Rapidly increasing cumulative incidence of coronavirus disease (COVID-19) in the European Union/European Economic Area and the United Kingdom, 1 January to 15 March 2020. Euro Surveill 2020.

4. Sun P, Lu X, Xu C, Sun W, Pan B: Understanding of COVID-19 based on current evidence. J Med Virol 2020.

5. Nakajima K, Kato H, Yamashiro T, Izumi T, Takeuchi I, Nakajima H, Utsunomiya D: COVID-19 pneumonia: infection control protocol inside computed tomography suites. Jpn J Radiol 2020.

6. Han R, Huang L, Jiang H, Dong J, Peng H, Zhang D: Early Clinical and CT Manifestations of Coronavirus Disease 2019 (COVID-19) Pneumonia. AJR Am J Roentgenol 2020: 1–6.

7. Dai WC, Zhang HW, Yu J, Xu HJ, Chen H, Luo SP, Zhang H, Liang LH, Wu XL, Lei Y et al: CT Imaging and Differential Diagnosis of COVID-19. Can Assoc Radiol J 2020: 846537120913033.

8. Chang H, Wen Q, Parvin B: Coupled Segmentation of Nuclear and Membrane-bound Macromolecules through Voting and Multiphase Level Set. Pattern Recognit 2015, 48(3):882–893.

9. Chan TF, Vese LA: Active contours without edges. IEEE Transactions on Image Processing 2001, 10(2):266–277.

10. Chang H, Zhou Y, Borowsky A, Barner K, Spellman P, Parvin B: Stacked Predictive Sparse Decomposition for Classification of Histology Sections. International Journal of Computer Vision 2014, 13(1):3–18.

11. Li M, Lei P, Zeng B, Li Z, Yu P, Fan B, Wang C, Li Z, Zhou J, Hu S et al: Coronavirus Disease (COVID-19): Spectrum of CT Findings and Temporal Progression of the Disease. Acad Radiol 2020.

12. Luo W, Yu H, Gou J, Li X, Sun Y, Li J, Liu L: Clinical Pathology of Critical Patient with Novel Coronavirus Pneumonia (COVID-19). Preprints 2020.

13. Paterlini M: Lockdown in Italy: personal stories of doing science during the COVID-19 quarantine. Nature 2020.

14. Saglietto A, D’Ascenzo F, Zoccai GB, De Ferrari GM: COVID-19 in Europe: the Italian lesson. Lancet 2020.

15. McMichael TM, Currie DW, Clark S, Pogosjans S, Kay M, Schwartz NG, Lewis J, Baer A, Kawakami V, Lukoff MD et al: Epidemiology of Covid-19 in a Long-Term Care Facility in King County, Washington. N Engl J Med 2020.

16. Takian A, Raoofi A, Kazempour-Ardebili S: COVID-19 battle during the toughest sanctions against Iran. Lancet 2020, 395(10229):1035–1036.

17. Kamerow D: Covid-19: the crisis of personal protective equipment in the US. BMJ 2020, 369:m1367.

18. Guan CS, Lv ZB, Yan S, Du YN, Chen H, Wei LG, Xie RM, Chen BD: Imaging Features of Coronavirus disease 2019 (COVID-19): Evaluation on Thin-Section CT. Acad Radiol 2020.

